# Impact of the cooked millet intervention on hemoglobin and other micronutrient deficiencies among anemic women of reproductive age

**DOI:** 10.1101/2025.11.22.25340788

**Authors:** Santosh Kumar Banjara, M.S. Radhika, Hemant Mahajan, R. Ananthan, Sourav Sen Gupta, Karthikeyan Ramanujam, J.J. Babu Geddam, Aruna vancha Reddy, G Vijayalakshmi, Ruchika Sharma, Swarna Lakshmi Janga, B.R. Nikhita, Devraj J Parasannanavar

## Abstract

**Background:** Iron deficiency Anemia and other micronutrient deficiencies among women of reproductive age is a significant public health concern in India with the prevalence exceeding 50%. The inclusion of micronutrient-rich millets in their habitual diet presents a sustainable strategy to improve micronutrient intake. This study aimed to evaluate the impact of millet-based recipes on hemoglobin levels and the overall micronutrient status of anemic women of reproductive age residing in Hyderabad, India.

**Methods:** A cluster-randomized controlled trial was conducted on 822 women aged 17 to 22 years (diagnosed with mild to moderate anemia), enrolled in Government 6 welfare residential degree colleges. Randomisation was done at hostel level and not at participants level to limit complications. The intervention group(3 hostels) consumed millet-based meals incorporating pearl millet, finger millet, and foxtail millet for 4–5 months, while the control group (3 hostels) continued their regular rice and wheat based. Venous blood samples were collected and Hemoglobin levels were assessed at three time points using a hematology autoanalyzer. Ferritin, Folate, Vit B-12,STFR, and Hs-CRP, levels were analyzed at both baseline and endpoint using the Abbott i-STAT 1000 autoanalyzer, Micronutrients (serum calcium and zinc) were analysed using AAS.

**Results:** The baseline characteristics were comparable between the two groups. The mean duration of intervention was 128 ± 11 days and the average millet intake was 50 g/day. Mean hemoglobin levels showed a significant improvement in the intervention group (10.40 ± 1.1 to 10.50 ± 1.46 g/dL), whereas a decline was observed in the control group (10.50 ± 1.06 to 10.30 ± 1.35), resulting in a statistically significant difference between the groups. The trajectory of mean hemoglobin levels over time demonstrated a significantly favorable trend in the intervention group compared to the control group (p <0.044). Moreover, the mean changes in folate and vitamin B12 levels from baseline to endpoint were more pronounced in the intervention group compared to the control group. Furthermore, there was a significant mean change observed across both groups. However, changes in other micronutrients such as ferritin, zinc, calcium, vitamin D, were comparable between the two groups from baseline to endline.

**Conclusions:** The Millet-based dietary intervention significantly improved hemoglobin, folate, and vitamin B12 levels among anemic women of reproductive age. However, no substantial changes were observed in other micronutrients. These findings highlight the potential of millets as a sustainable dietary strategy for anemia management. Further research is needed to explore long-term benefits and integration into public health programs.

## INTRODUCTION

Anemia and micronutrient deficiencies remain significant public health challenges among women of reproductive age, particularly in low- and middle-income countries. According to the World Health Organization (WHO), approximately 29.9% of women aged 15–49 years were anemic in 2019, affecting over half a billion women globally (*WHO, 2021*). In India, the prevalence is alarmingly high, with the National Family Health Survey-5 (NFHS-5) reporting that 57% of women of reproductive age suffer from anemia (*IIPS & ICF, 2021*). Iron deficiency remains the leading cause, but other micronutrient deficiencies, including folate, vitamin B12, vitamin D, and zinc, also contribute to poor health outcomes such as fatigue, cognitive impairment, weakened immunity, and adverse pregnancy outcomes (*Ward 2014;* Bhaskaram, 2001). Optimal hematopoietic function requires adequate levels of folate and vitamin B12, which are vital for central nervous system metabolism. These nutrients play a crucial role in methylation reactions essential for neurotransmitter, phospholipid, and nucleotide synthesis (*Fishman, et.al, 2000; Chanarin, et.al, 1989; Penninx, et.al, 2000*)

The etiology of anemia is multifactorial, encompassing inadequate dietary intake, poor nutrient bioavailability, and increased physiological demands, which are particularly pronounced in adolescent girls and young women (*Thankachan, et.al, 2007*). Traditional staple cereals such as rice and wheat, commonly consumed in India, are low in iron and other essential micronutrients, further exacerbating the issue (*Kumar et al., 2022*). In this context, millet consumption has gained attention as a viable dietary intervention to combat anemia and micronutrient deficiencies (Givens *et al*., 2024).

Millets are highly nutritious cereals rich in iron, zinc, calcium, and dietary fiber, with a low glycemic index and superior micronutrient bioavailability compared to conventional grains (*Anitha et al., 2021*). Among various millet species, **finger millet** (*Eleusine coracana*), **foxtail millet** (*Setaria italica*), and **pearl millet** (*Pennisetum glaucum*) have been particularly recognized for their health benefits. Finger millet is one of the richest plant-based sources of calcium and also contains significant amounts of iron and dietary fiber, making it beneficial for improving hemoglobin levels (*Moharana, et al., 2020*). Foxtail millet is a good source of iron, protein, and antioxidants and has been found to enhance iron bioavailability and improve metabolic health (*Sharma, et.al, 2018*). Pearl millet is notably high in iron and zinc and has demonstrated efficacy in improving iron status and reducing anemia prevalence among at-risk populations (*Manwaring, et al., 2016*).

Despite the known nutritional advantages of millets, there is limited evidence from controlled dietary intervention studies evaluating their long-term impact on hemoglobin and other micronutrient levels in anemic women. This study aims to substitute traditional rice-and wheat-based meals with selected cooked millet recipes and assess cooked millet-based dietary intervention incorporating **finger millet, foxtail millet, and pearl millet** on hemoglobin and key micronutrient status, including iron, folate, vitamin B12, zinc, calcium, and vitamin D, among anemic women aged 17–22 years. The findings will contribute to the growing body of evidence on millet-based dietary interventions and offer insights into effective public health strategies to combat anemia and micronutrient deficiencies.

Iron and folic acid (IFA) supplements may cause side effects like nausea, stomach pain, bloating, constipation, abdominal discomfort, and dark stools (*Ramya, 2016*). While IFA supplementation can improve hemoglobin and folate levels in cases of iron deficiency anemia, it may not be as effective if deficiencies in other micronutrients cause anemia. Hence, this study aims to address anemia by targeting both hemoglobin levels and other essential micronutrients through a comprehensive approach. Integrating programs and utilizing a sustainable crop to address anemia in adolescent girls also contributes to the achievement of the Sustainable Development Goals, particularly 2, 5, 12, 13, and 17.

### Primary objectives

To determine the efficacy of millet-based composite diets (Pearl millet, Finger millet, Foxtail millet) on hemoglobin status by replacing one meal (breakfast/ lunch) among anemic women of reproductive age

### Secondary objective

To determine the efficacy of a millet-based diet on micronutrient status and body composition by replacing one meal among anemic women of reproductive age.

## MATERIALS AND METHODS

**Design:** A **Randomized Control Trial (RCT)** was designed for a six-month parallel-group intervention. Healthy women of reproductive age (17-22 years) with mild and moderate anemia.

**Site of the study:** The study was conducted in hostels under the Social and Tribal Welfare Department in and around Hyderabad. Residential hostellers were chosen over day scholars to ensure uniform dietary intake, as all meals were prepared in a common kitchen. Approval for the study was granted in writing by the Social and Tribal Welfare Department of the Government of Telangana. Additionally, meetings were held with hostel authorities, students, parents, and institute chiefs to explain the study’s objectives, procedures, and potential risks. Informed written consent was obtained from participants and their parents before initiating the intervention.

**Ethical consent:** Ethical approval was secured from the Institutional Ethics Committee (Protocol No. 02/I/2022 dated 11/02/2022, RR/1/V/2022 dated 09/11/22, R/CR/2I/2023 dated 03/04/2023). The study was also registered with the Clinical Trial Registry of India (CTRI: REF/2022/06/055578). Permission was obtained from the Social Welfare Department, Govt. of Telangana.

**Sampling strategy:** Based on the previous studies, a sample size of 812 individuals was needed to detect a mean difference of 0.6g/dl hemoglobin level with a standard deviation of 1.5, with 80% power and 5% significance level and 20% attrition rate with a design effect of 3.45. It is provided in Table 1. The sample size required per group was 406.

**Table 1.**
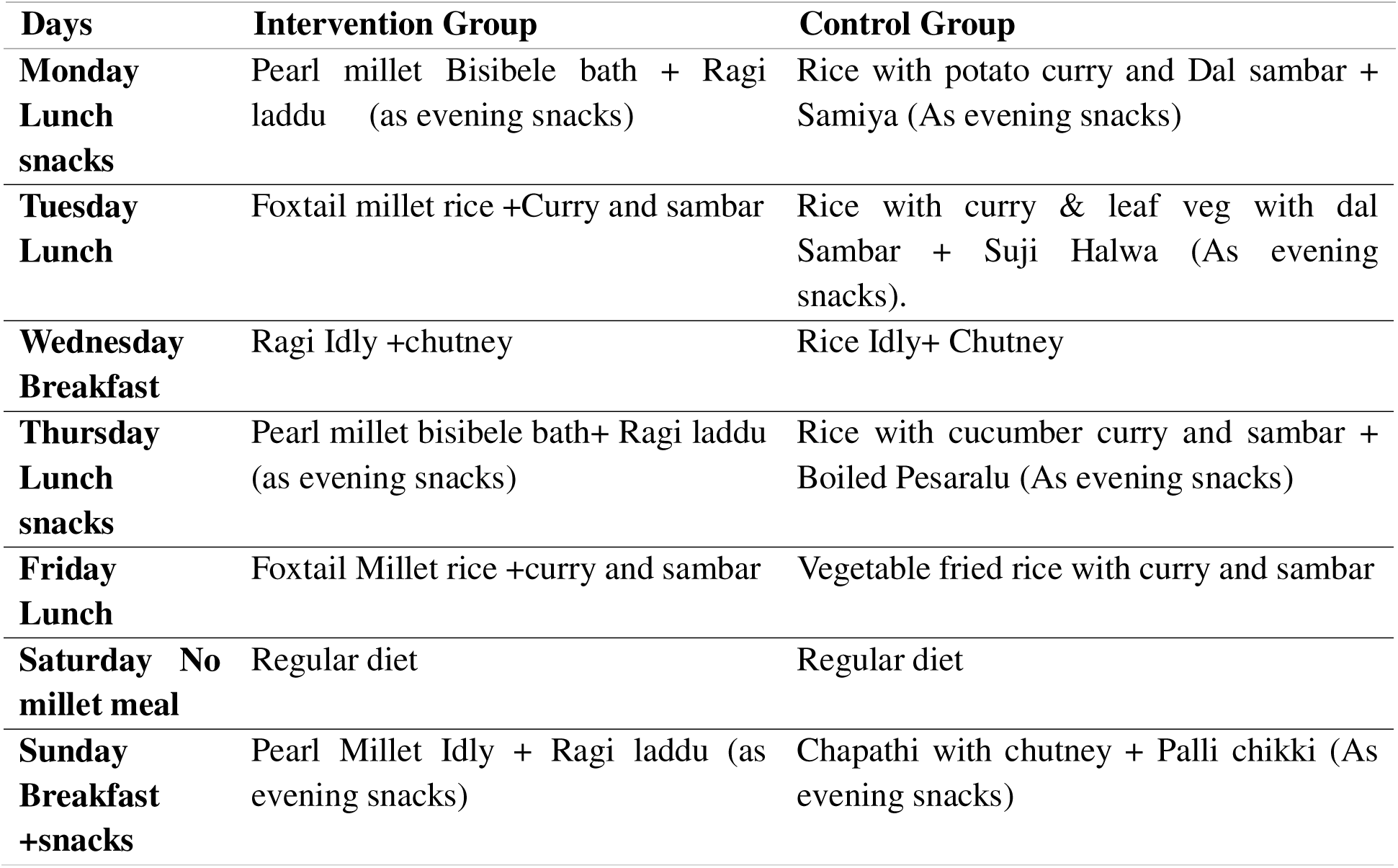
Weekly diet menu for intervention and control group.

| Days | Intervention Group | Control Group |
| --- | --- | --- |
| **Monday Lunch snacks** | Pearl millet Bisibele bath + Ragi laddu (as evening snacks) | Rice with potato curry and Dal sambar + Samiya (As evening snacks) |
| **Tuesday Lunch** | Foxtail millet rice +Curry and sambar | Rice with curry & leaf veg with dal Sambar + Suji Halwa (As evening snacks). |
| **Wednesday Breakfast** | Ragi Idly +chutney | Rice Idly+ Chutney |
| **Thursday Lunch snacks** | Pearl millet bisibele bath+ Ragi laddu (as evening snacks) | Rice with cucumber curry and sambar + Boiled Pesaralu (As evening snacks) |
| **Friday Lunch** | Foxtail Millet rice +curry and sambar | Vegetable fried rice with curry and sambar |
| **Saturday No millet meal** | Regular diet | Regular diet |
| **Sunday Breakfast +snacks** | Pearl Millet Idly + Ragi laddu (as evening snacks) | Chapathi with chutney + Palli chikki (As evening snacks) |

### Selection criteria

**Inclusion criteria:** The study included healthy women of reproductive age (17–22 years) who were identified as having mild (Hb: 11–11.9 g/dL) or moderate (Hb: 8.0–10.9 g/dL) anemia. Participants were required to provide informed consent before enrolment in the study.

**Exclusion criteria:** Women who were undergoing treatment for thyroid disorders, polycystic ovarian disease (PCOD), or other chronic conditions were excluded from the study. Additionally, individuals with genetic blood disorders such as thalassemia and sickle cell anemia, as well as those diagnosed with hemochromatosis, chronic diarrhea, or chronic gastric conditions (including inflammatory bowel disease, Crohn’s disease, gastric ulcers, celiac disease, and hemorrhagic disorders), were not eligible for participation. Furthermore, participants with severe anemia (Hb < 8.0 g/dL) were excluded and referred for medical care. Women with normal hemoglobin levels (≥12 g/dL) and those already enrolled in any ongoing clinical trials were also not considered for the study.

### Health Screening

Participants were screened for pre-existing conditions, including chronic diseases, gastrointestinal disorders, and severe anemia. Only those meeting the inclusion criteria were enrolled in the study for further procedures. The selected participants were randomized.

### Randomization

It was done at the hostel level, not at the participant’s, to minimize the complications of feeding two different dietary interventions in a single hostel. Other confounding variables, such as curries, which were common among both groups, were kept to a minimum. 821 subjects aged between 17 and 22 were recruited for the study sample. 409 subjects were chosen for a millet-based meal (intervention group) supplementation in 3 hostels, and the remaining (n=413) were chosen from other hostels for the control group (Regular diet). Following randomization, participants proceeded with further assessments.

### Baseline Assessment

The baseline assessment involved evaluating the participant’s **Socio-Demographic,** anthropometric, hematological, biochemical, and dietary parameters before initiating the dietary intervention. This ensured homogeneity between the intervention and control groups and provided a reference point for measuring changes post-intervention.

**Anthropometric Measurements: Height:** Measured using a SECA 213 calibrated stadiometer, recorded to the nearest 0.5 cm.

**Weight:** Measured using a digital electronic weighing scale, recorded to the nearest 0.1 kg.

**Body Mass Index (BMI):** Calculated using the standard formula and classified according to WHO guidelines.

**Body Composition:** Assessed using the InBody-121 body composition analyzer, which operates on the bioelectrical impedance analysis (BIA) principle to measure body fat, muscle mass, and water content.

#### i. Biochemical Parameters

Venous blood samples (10ml/person) were collected in EDTA vacutainers and analyzed using the following instruments and kits:

**Complete Blood Count (CBC):** Measured using the Horiba ABX Micros ES 60 hematology autoanalyzer, which provided quantitative data on hemoglobin (Hb), red blood cell indices, and platelet count. Calibration was performed daily using Horiba Minotrol 16 reference standards (high, normal, and low levels) (reference of Dr.Ravindernath Paper)

**Serum Ferritin:** Analyzed using CMIA technology on the Abbott Architect System with the ARCHITECT Ferritin Kit (7K59) (*Bhargava, et.al, 2020*).

**Vitamin D (25-OH D):** Measured using CLIA on the Abbott Architect autoanalyzer (ARCHITECT 25-OH Vit-D 5P02) (Kokkinari, et.al, 2024)

**Folate and Vitamin B12:** Determined using CMIA on the Abbott Architect autoanalyzer with ARCHITECT Folate Kit (1P74) and B12 Kit (7K61), respectively (*Shivkar, et.al, 2022*).

### Thyroid Function Tests (TFTs)

**Total T3, Total T4, and TSH:** Measured using CMIA technology on the Abbott Architect iSystem with specific reagent kits (ARCHITECT TSH (7K62), Total T3 (7K64) and Total T4 (7K66)) (*Dhillon-Smith, et.al, 2019*).

**Soluble Transferrin Receptor (sTfR):** Measured using the Cobas autoanalyzer through chemiluminescent immunoassay (CLIA).

**Inflammation Marker: High-Sensitivity C-Reactive Protein (hs-CRP):** Quantified using immunoturbidimetric assay on the Cobas autoanalyzer.

**Micronutrient Levels:** Serum zinc and calcium levels were measured using Atomic Absorption Spectroscopy (AAS) at wavelengths of 213.9 nm and 422.7 nm, respectively.

### Dietary intake

Institutional dietary data were collected from participants to evaluate their dietary intake patterns and nutrient consumption over three days. Socio-demographic data, including age, education, economic status, and lifestyle factors, were collected.

### Dietary Intervention

The dietary intervention was designed for a six-months duration. The weekly diet menu for both the intervention and control groups is presented in Table 2. Participants in the intervention group received cooked millet-based recipes, while those in the control group continued with their regular rice and wheat-based diets. Cooks were trained and closely supervised in millet recipe preparation until they gained proficiency. To promote good hygiene practices and minimize food wastage, teachers and nutrition experts conducted interactive sessions on personal hygiene and the nutritional benefits of millet. These sessions were held after the sensory evaluation and during the feeding program, using focus groups and educational posters. Participants were monitored by teachers and project staff to ensure proper handwashing before meals and to prevent food wastage. Regular feedback on the millet-based diets was encouraged and used to refine recipes throughout the study to enhance palatability.

**Table 2.** Baseline Sociodemographic, anthropometric, anemia prevalence and mean intake of nutrients of both the group.

| Parameters | Intervention group | Control group |
| --- | --- | --- |
| No of total students in the hostels (3 in each) | 1210 | 1554 |
| No of the subjects screened | 1147 | 1472 |
| Long Absentees | 63 | 82 |
| No of the subjects were haematologically screened | 811 | 746 |
| No of Subjects $\geq 12$ g/dl- normal | 345 (42.54%) | 307 (41.2%) |
| No mild and moderate anemic subjects | 409 (50.43%) | 417 (55.9%) |
| No of Severe - $<8.0$ gm/dl N+ % | 57 (7.02%) | 22 (2.9%) |
| No of the Subjects enrolled for intervention | 409 | 413 |
| Dropouts/ Not interested to participate | 0 | 4 |
| Prevalence of anemia | 57.45% | 58.8% |
| **Demographic details of included subjects** |  |  |
| Mean Age | 19 | 19.4 |
| Mean Weight | 46.69 ± 7.78 | 46.37 ± 7.41 |
| Mean Height | 152.50 ± 5.52 | 152.44 ± 5.72 |
| Mean BMI | 20 ± 2.9 | 19.9 ± 2.9 |
| **Mean Nutrient intake among intervention and control participants at baseline** |  |  |
| Nutrients | Intervention | Control |
| Energy (kcal) | 1292.38 ± 64 | 1312.41 ± 33 |
| Carbohydrate (g) | 199.238 ± 24 | 226.41 ± 24 |
| Protein (g) | 47.62 ± 3 | 44.23 ± 5 |
| Fat (g) | 45.13 ± 1 | 45.10 ± 15 |
| Dietary fiber (g) | 28.88 ± 1 | 24.17 ± 2 |
| **Calcium (mg)** | 509.82 ± 14 | 605.19 ± 69 |
| **Iron(mg)** | 15.92 ± 0.6 | 15.26 ± 1 |
| **Total folate (µg)** | 253.45 ± 6 | 215.03 ± 21 |
| **Vitamin C** | 69.70 ± 12 | 57.47 ± 7 |

### Mid-point analysis

At the study’s midpoint, hemoglobin levels and body composition were assessed in participants from both groups.

### Endpoint analysis

At the endpoint of the study, biochemical parameters were assessed to evaluate the impact of the intervention. Hemoglobin levels were measured to determine changes in anemia status, while soluble transferrin receptor (sTfR) and serum ferritin levels were analyzed to assess iron status. Essential micronutrients, including serum zinc, calcium, vitamin D, folate, and vitamin B12, were evaluated to monitor nutritional improvements. Inflammatory markers such as high-sensitivity C-reactive protein (hs-CRP) were also measured. Additionally, thyroid function tests, including total T3, total T4, and thyroid-stimulating hormone (TSH), were conducted. All assessments were performed using standardized laboratory procedures, ensuring the accuracy and reliability of the results.

### Statistical analysis

Data collection was conducted using a pre-tested proforma and recorded in an Excel sheet. A diet chart was provided to the study supervisor to monitor compliance. Statistical analyses were performed using SPSS version 19. For normally distributed variables, descriptive statistics were presented as means and standard deviations. A paired t-test was used to analyze continuous variables within the groups, while the Wilcoxon rank-sum test was applied for comparisons between groups. A p-value of less than 0.05 was considered statistically significant for all analyses. The recruited and analyzed participants are presented in the CONSORT flowchart. The overall flowchart of the study is given in Figure 1.

**Figure 1.**
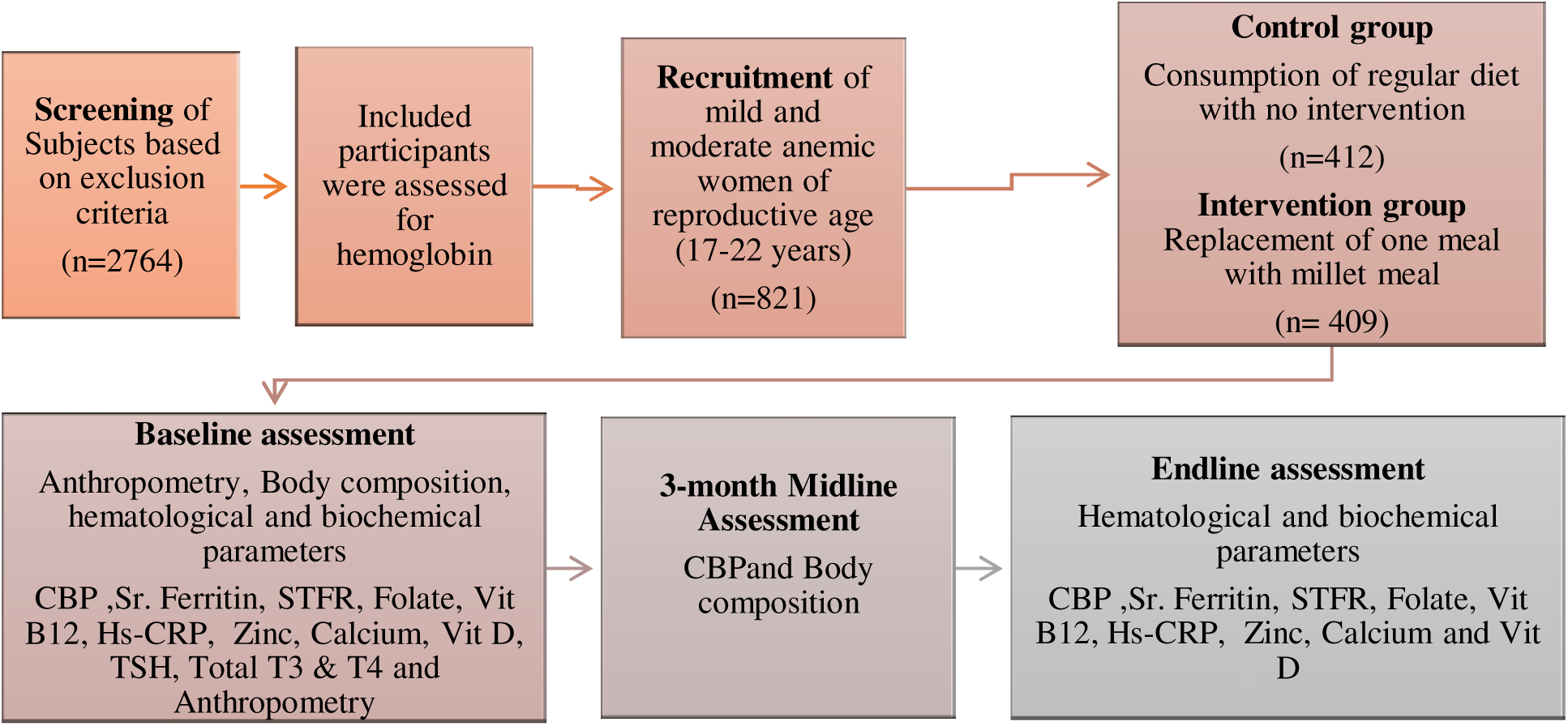
Overall flowchart of the study.

**Figure 2.**
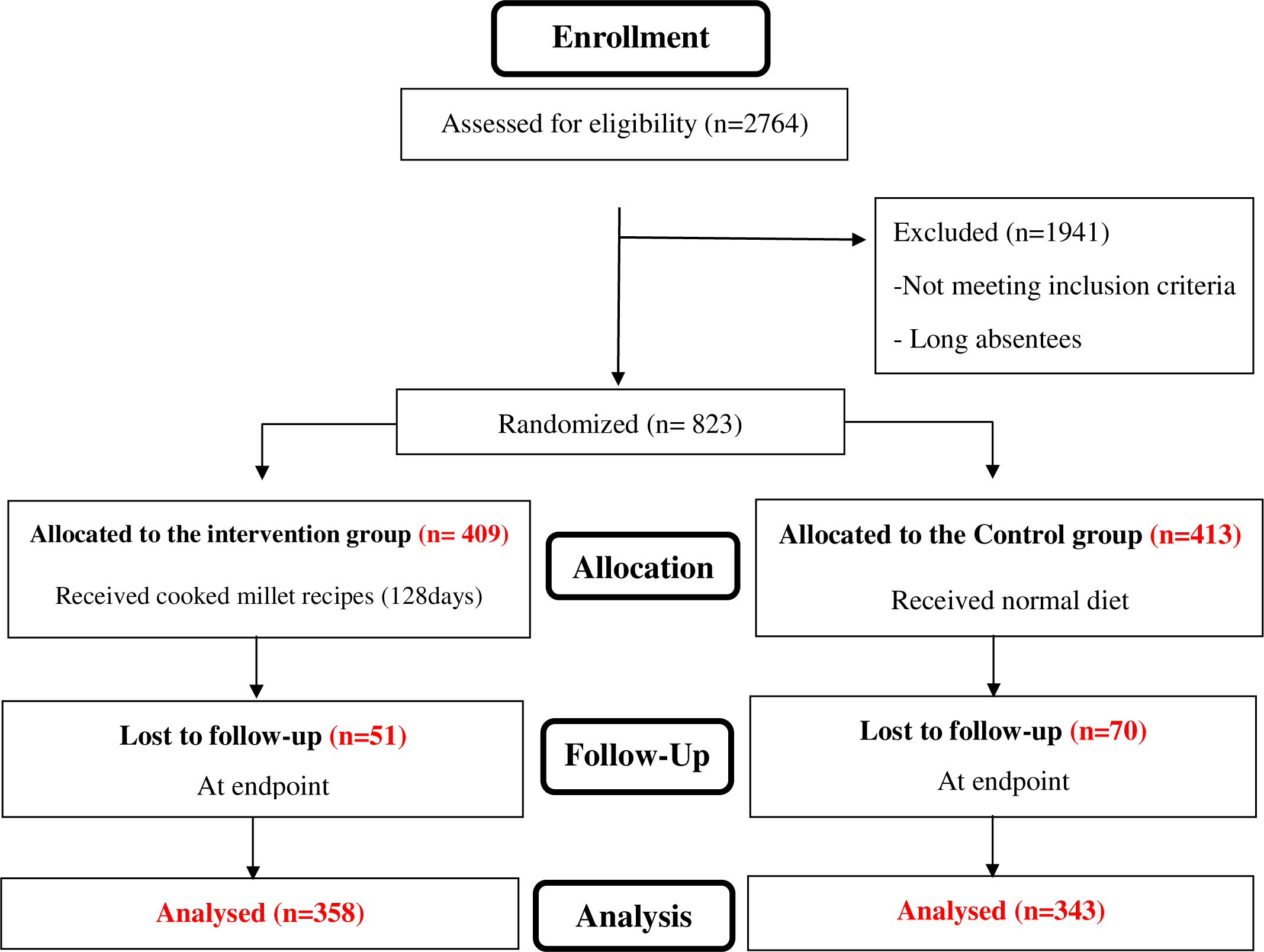
CONSORT Flowchart of the study.

**Figure 3.**
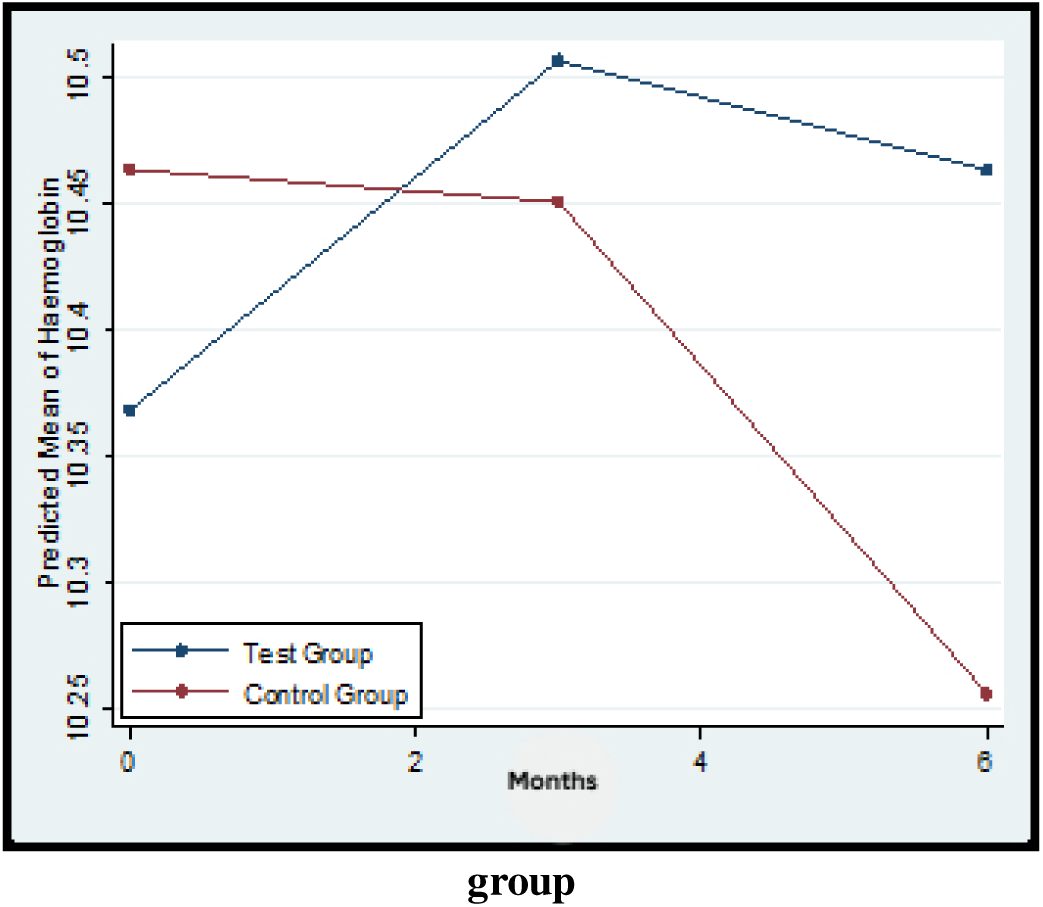
Graphical representation of mean haemoglobin at 3 time points among both the group. Predicted mean haemoglobin levels over 6 months in the test and control groups.. The test group demonstrates an initial rise followed by a modest decline, whereas the control group shows a gradual decrease across the observation period.

## Results

Baseline data in table 2 highlights that a total of 1147 subjects screened for the intervention group and 1472 for the control group. From these, 409 and 413 participants were enrolled in the study, respectively. As presented in table 3, the demographic characteristics of both groups were comparable. The mean intake of most nutrients showed little difference prior to the start of the intervention, except that the intervention group had a slightly higher intake of protein and fiber, while the control group consumed more carbohydrates. The mean hemoglobin level was marginally higher in the control group (10.50 g/dL ± 1.1) compared to the intervention group (10.40 g/dL ± 1.06). Additionally, the levels of micronutrients and vitamins were found to be nearly equivalent in both groups before the intervention. The duration of interventions varied between 115 and 136 days across the three hostels, with one meal substituted by one of these 8 recipes.

**Table 3.**
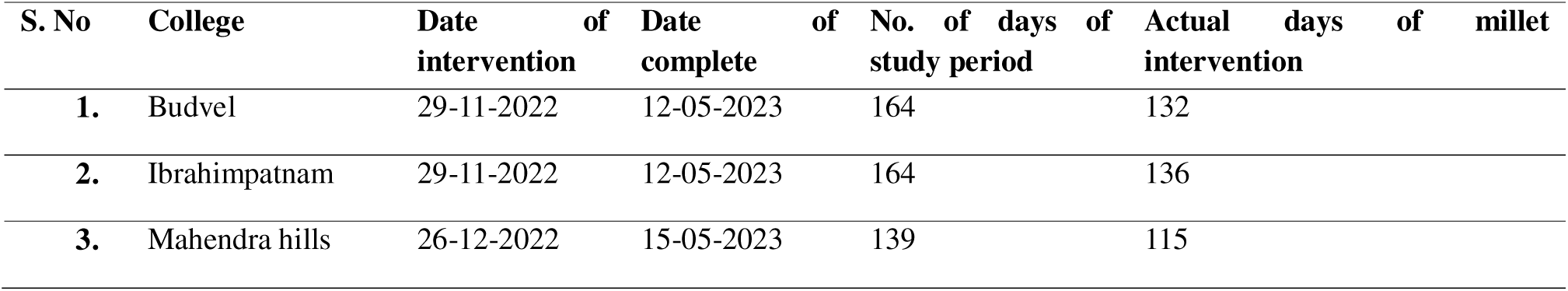
Duration of Institutional-wise Recipe Consumption.

### Effect of intervention on Heamoglobin, Micronutrients and vitamin status

Tables 4 and 5 collectively illustrate the impact of the millet-based dietary intervention on micronutrient levels and anemia within the study group. The results from both average figures and percentages indicate consistent trends, with significant enhancements in various hematological and micronutrient measures. The trajectory of mean levels of hemoglobin differs over time between two groups (p-value = 0.0436). In Table 4, hemoglobin concentrations slightly increased in the intervention group, while the control group experienced a slight decrease of (p=0.003) This is corroborated by Table 5, which shows the proportions of 15.4% at the endpoint in the intervention group, in contrast to to 13.4% in the control group. The prevalence of moderate anemia decreased from 60.5% to 54.3% in the intervention group, while it saw a slight rise in the controls. Prevalence of IDA reduced by 15.22% in the intervention group and by 12.34% in the control group, indicating a 2.88% higher reduction in IDA among participants receiving the millet-based intervention. Both folate and vitamin B12 levels exhibited significant improvements in the intervention group (+2.5 ng/mL and +49 pg/mL respectively), along with corresponding increases in the percentage of children falling into the normal range. Folate deficiency dropped from 95.1% to 82.9%, and vitamin B12 deficiency decreased from 79.3% to 60.9%. In contrast, the control group experienced a slight increase in folate deficiency with vitamin B12 deficiency remaining high, highlighting the specific advantages of the intervention. The occurrence of ferritin deficiency fell from 93.6% to 89.5% in the intervention group, while the control group also demonstrated some improvement. However, not all indicators showed favorable changes. Zinc levels dropped in both groups, with a more significant rise in zinc deficiency in the intervention group (6.3% to 24.7%). Vitamin D deficiency persisted widely, with over 90% still lacking sufficient levels at the endpoint. Calcium levels experienced modest improvements in both groups, and hs-CRP trends indicated no significant alteration in inflammation status. No significant within-group changes were observed in body fat mass, skeletal fat mass, or percent body fat in either group. However, at midpoint, the intervention group showed significantly higher skeletal fat mass (*p* = 0.04). No major adverse events were reported due to consumption of millet-based recipes.

**Table 4.**
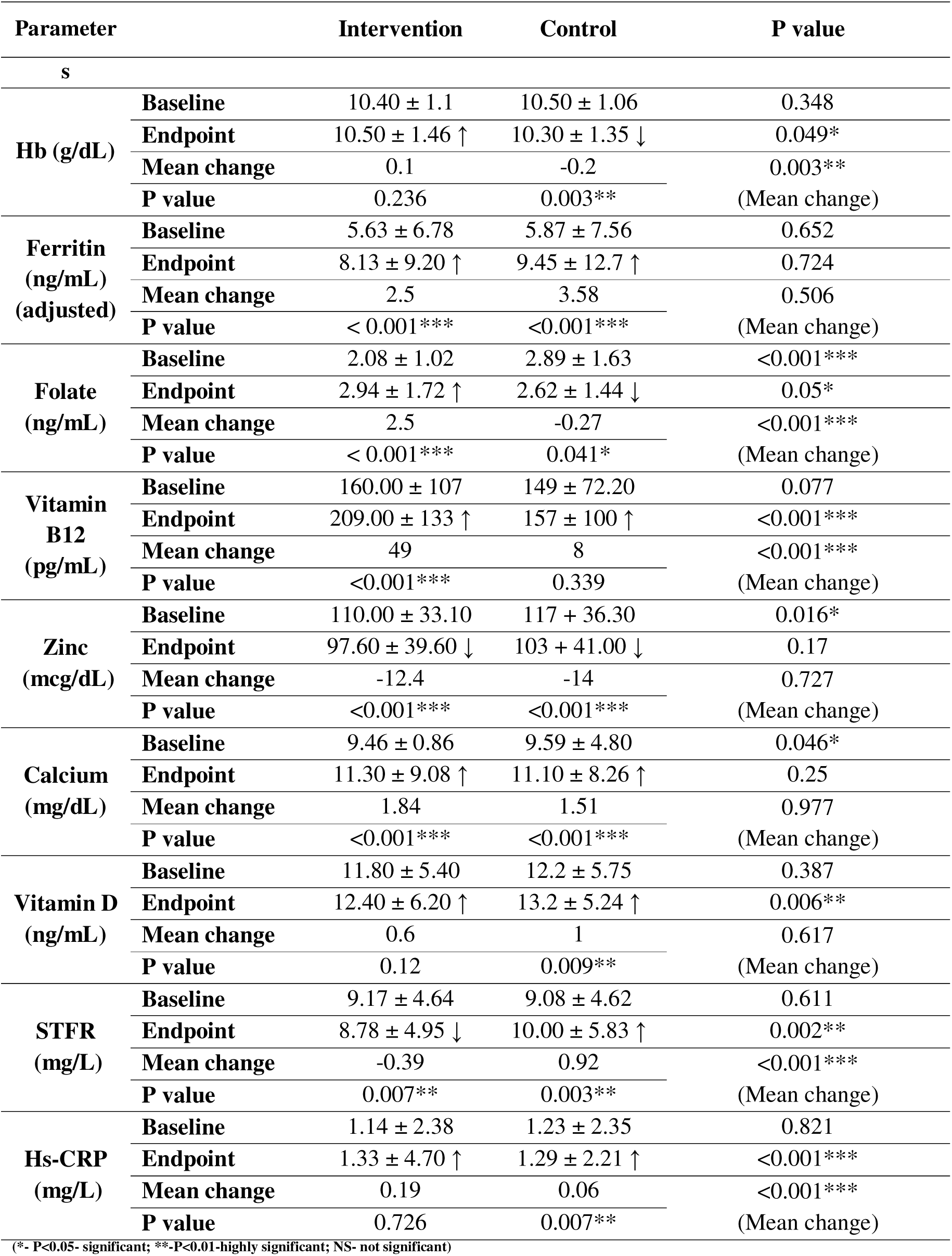
Effect on mean values of hemoglobin and micronutrients of both groups.

**Table 5.** Effect of millet on proportions of anemia and micronutrient deficiencies.

| Biochemical | Category | Intervention n (%) | Control n (%) |
| --- | --- | --- | --- |

| parameters |  | Baseline | Endpoint | Baseline | Endpoint |
| --- | --- | --- | --- | --- | --- |
| **Anemia (Hb)** | Normal | 0 (0.00%) | 55 (15.40%) | 0 (0%) | 46 (13.41%) |
| (Cut-offs: Normal: | Mild | 161 (39.46%) | 94 (26.33%) | 165 (39.95%) | 61 (17.78%) |
| ≥12 g/dL; Mild: | Moderate | 247 (60.53%) | 194 (54.34%) | 248 (60.05%) | 218 (63.56%) |
| 11-11.9 g/dL; | Severe | 0 (0.00%) | 14 (3.92%) | 0 (0%) | 18 (5.25%) |
| Moderate: 8-10.9 g/dL; Severe: ≤7.9 g/dL | **Total** | 408 (100%) | 357 (100%) | 413 (100%) | 343 (100%) |
| **Ferritin adjusted** | Deficient | 366 (93.60%) | 307 (89.50%) | 378 (93.10%) | 276(87.61%) |
| Adjusted ferritin | Normal | 25 (6.39%) | 36 (10.49%) | 28 (6.89%) | 39 (12.38%) |
| <15 ng/mL | **Total** | 391 (100%) | 343 (100%) | 406 (100%) | 315 (100%) |
| **Folate** | Deficient | 372 (95.14%) | 292 (82.95%) | 340 (82.92%) | 281 (88.36%) |
| Deficiency- Folate | Normal | 19 (4.85%) | 60 (17.04%) | 70 (17.10%) | 37 (11.63%) |
| < 4 ng/mL | **Total** | 391 (100%) | 352 (100%) | 410 (100%) | 318 (100%) |
| **Vitamin B<sub>12</sub>** | Deficient | 310 (79.28%) | 215 (60.91%) | 336 (81.95%) | 258 (81.13%) |
| Deficiency < 203 pg/mL | Normal | 81 (20.72%) | 138 (39.09%) | 74 (18.04%) | 60 (18.86%) |
|  | **Total** | 391 (100%) | 353 (100%) | 410 (100%) | 318 (100%) |
| **Zinc** | Deficient | 20 (6.29%) | 82 (24.70%) | 19 (5.97%) | 74 (23.19%) |
| Deficiency: Zn ≤ 70 mcg/dL | Normal | 298 (93.71%) | 251 (75.30%) | 299 (94.02%) | 253 (76.81%) |
|  | **Total** | 318 (100%) | 333 (100%) | 318 (100%) | 318 (100%) |
| **Calcium** | Deficient | 70 (10.06%) | 58 (19.72%) | 85 (27.07%) | 65 (22.96%) |
| Deficiency: Ca ≤ 9 mg/dL | Normal | 243 (89.94%) | 236 (80.27%) | 229 (72.93%) | 218 (77.03%) |
|  | **Total** | 313 (100%) | 294 (100%) | 314 (100%) | 283 (100%) |
| **Vitamin D** | Deficient | 369 (92.71%) | 330 (94.28%) | 382 (93.17%) | 292 (91.82%) |
| Deficiency: Vit D < 20 ng/mL | Insufficient | 29 (7.28%) | 17 (4.85%) | 28 (6.82%) | 26 (8.17%) |
| Insufficiency: 20–40 ng/mL | Normal | 0 (0%) | 3 (0.85%) | 0 (0%) | 0 (0%) |
|  | **Total** | 398 (100%) | 350 (100%) | 410 (100%) | 318 (100%) |
| **sTFR** | Elevated | 384 (98.20%) | 340 (97.98%) | 396 (97.53%) | 308 (97.78%) |
| Elevated sTFR >2.97mg/L) | Normal | 7 (1.79%) | 7 (2.05%) | 10 (2.46%) | 7 (2.23%) |
| Normal Stfr ≤2.97mg/L) | **Total** | 391 (100%) | 347 (100%) | 406 (100%) | 315 (100%) |
| **HsCRP** | Elevated | 20 (5.12%) | 17 (4.95%) | 23 (5.66%) | 11 (3.49%) |
| Elevated - HsCRP>5mg/L) | Normal | 371 (94.88%) | 326 (95.04%) | 383 (94.33%) | 304 (96.50%) |
| Normal - HsCRP≤5mg/L) | **Total** | 391 (100%) | 343 (100%) | 406 (100%) | 315 (100%) |

## Discussion

The rate of anemia among adolescents in India stands at 58.9% (NFHS-5). It continues to affect the productivity and health of the population, especially in low- and middle-income countries. Anemia has lot of physiological implications that adversely affects the patient; Adolescent girls who suffer from anemia often remain anemic during their pregnancies, increasing the likelihood of maternal mortality, premature births, low birth weights, and infant deaths, all this and more ultimately resulting in a 1.2% loss in India’s GDP (Chakrabarty et al., 2023). 44.1% anemic pregnant women consumed IFA tabelts during their last pregnancy and coverage of for IFA tablets among adolescent women was 28 %; IFA uptake among women and children has been low for several reasons (Rai *et al*., 2023). Anemia has been a prominent public health concern in India for many years, and tackling it requires behavioral changes, creating supportive environments, and raising awareness for long-term effectiveness. Employing a range of millet types instead of focusing solely on one, making it more feasible for participants to adhere to the intervention. Growing evidence in the existing literature suggests that improving food environments or promoting favorable conditions can have a substantial impact on the effectiveness of interventions; 14 observational studies have demonstrated a connection between food environment characteristics and dietary, nutritional, and health outcomes (Westbury et al.,2021).

### Effect of intervention on Heamoglobin, micronutrients and vitamin

Recent observations from the intervention indicate positive but modest improvements in children’s hematological status. In another study by Scott et al., (2019) School-going children aged 12–16 years who received the six-month fortified pearl millet dietary intervention showed a small rise in hemoglobin of about 0.1 g/dL. Although modest, this improvement is consistent with previous research demonstrating the benefits of millet consumption. Another study reported meaningful increases in hemoglobin and ferritin levels among adolescents consuming finger-millet or pearl-millet foods. Together, these findings suggest that regularly eating millet can support gradual improvements in iron status among children. (Karkada et al., 2019)

Millets are not a rich source of folate and vitamin B12, yet the adolescent girls observed an increase after the intervention, likely due to their high fiber content (Dong et al., 2019). This fiber aids in enhancing gut microbiota, and the microbial communities in the human gut can synthesize both vitamins through aerobic and anaerobic metabolic pathways to meet their own needs, regardless of the initial levels of vitamin B12 and folate present (Cronin et al.,2021; Fang et al., 2017; Grant et al., 2024).

An increase in calcium levels was noted in both groups, which may be attributed to the high calcium content in millets, and these findings align with the research conducted by (singh et al., 2020).

No significant rise was seen in zinc and Vitamin D among both the groups after intervention, due to cooking and soaking zinc tends to leach out and also bioaccessibility of zinc is reduced (Gowda et al., 2022; Hemalatha et al., 2007). Vit D is usually low in millets and might be the reason for persistent deficiency even after intervention (Jacob et al., 2024).

A systematic review on Mediterranean diet and its effect coupled with other intervention concluded that alone dietary intervention might not be effective in changing body composition, to see a significant changes multi-pronged approach is best suited, hence no significant changes were seen in the present study that only included one meal replacement without any other invention (sood et al., 2025)

### Implications

This research was an efficacy study carried out in a practical setting. Adolescent women given a composite meal featuring millet as a substitute for the commonly consumed wheat or rice. The findings of this research indicate that substituting one meal per day with a millet-based recipe may enhance the overall macro and micro nutrient intake along with diet diversity. These millet-based recipes can be integrated into the existing Saksham Anganwadi and POSHAN 2.0 programs for adolescent girls, which enrolled 2.2 million participants in the year 2022-2023. Already initiative from government and non-profit organisation to promote consumption and inclusion of millet as ready to eat porridge mix and take home ration are going on in different parts of the India, this can be a good inclusion for the same. This approach would be more sustainable, culturally relevant, and comprehensive than the current iron and folic acid (IFA) supplementation as compliance is the major drawback. In addition to addressing anemia, millets are linked to a beneficial phytochemical profile and better gut health (Anitha *et al,* 2024).

### Strengths

One of the key strengths of this study is its attempt to fill the gap in existing interventions focused on IFA tablet supplementation, which primarily addresses iron and folate deficiency anemia, which overlooks importance of other nutrients such as B12 that may contribute to anemia. Standard National Iron Folic Acid supplementation has low compliance, coverage and has shown to alter gut microbiota leading to symptoms like constipation and bloating (Bloor *et al.,*2021). Implementing a food-based intervention to combat anemia also promotes sustainable food systems. Incorporating millet into familiar recipes that usually contain rice or wheat, along with offering various options, has fostered an enabling environment that leads to improved nutritional outcomes. Another aspect of the study was designed to create a setting that encourages behavioral change among children, which included integrating traditional millets consumed by the local population, as well as using three different varieties instead of just one, as seen in previous research; this approach enhances feasibility for participants in adhering to the intervention.

### Limitations

The intervention was intended to last for 180 days; however, due to an unexpected academic break, the full duration of the intervention was not completed. Recent meta-analysis have suggested the longer the duration the better the outcome with millet supplementation. A multi-faceted approach may be more effective for nutrients such as zinc and vitamin D, as well as for observing noticeable changes in the body composition of the participants.

## Conclusion

The research suggests that replacing a typical meal of rice or wheat with one that includes millets may enhance hemoglobin, ferritin, folate, and vitamin B12 levels in anemic adolescent women. This intervention represents a more sustainable and holistic strategy that can be integrated into the already existing welfare programs like SAG and POSHAN 2.0. Which reaches millions of anemic girls in India.

## Informed Consent Statement

All participants provided consent after being informed about the study purpose, procedures, and their right to withdraw at any time. Confidentiality was maintained throughout the study.

## CRediT authorship contribution statement

Santosh Kumar Banjara: Writing- Original Draft, Writing-Review and editing

Supervision, Visualisation, Investigation, Project Administration,

Radhika M.S: Writing- Review and Editing, Validation, Methodology

Hemant Mahajan: Software, data Curation, Formal Analysis, Writing- Review and editing Sourav Sen Gupta: Visualization, Writing: Review & editing

R. Ananthan: Formal Analysis, Validation, Investigation, Writing – Review and Editing

J.J. Babu Geddam: Resources, Project Administration, Karthikeyan Ramanujan: Formal Analysis, Data Curation Vijaya Lakshmi G: Project Administration

Aruna Vancha Reddy: Project Administration

Swarana Lakshmi Janga: Data Curation and Project Administration

Ruchika Sharma: Data Curation

Devraj J Parasannanavar: Conceptualization, Supervision, Investigation, Visualisation,

Project Administration, Writing: Review and Editing

## Data Availability

All data produced in the present study are available upon reasonable request to the Corresponding author.

## Acknowledgement

The authors gratefully acknowledge the Director, ICMR-National Institute of Nutrition, Hyderabad, for providing the opportunity and necessary support to carry out this work. The authors declare that no funding was received for this study.

